# Overlap between COPD genetic association results and transcriptional quantitative trait loci

**DOI:** 10.1101/2024.07.08.24310079

**Authors:** Aabida Saferali, Wonji Kim, Robert P. Chase, NHLBI TransOmics in Precision Medicine (TOPMed), Chris Vollmers, Edwin K. Silverman, Michael H. Cho, Peter J. Castaldi, Craig P. Hersh

## Abstract

**Rationale:** Genome-wide association studies (GWAS) have identified multiple genetic loci associated with chronic obstructive pulmonary disease (COPD). When integrated with GWAS results, expression quantitative trait locus (eQTL) studies can provide insight into biological mechanisms involved in disease by identifying single nucleotide polymorphisms (SNPs) that contribute to whole gene expression. However, there are multiple genetically driven regulatory and isoform-specific effects which cannot be detected in traditional eQTL analyses. Here, we identify SNPs that are associated with alternative splicing (sQTL) in addition to eQTLs to identify novel functions for COPD associated genetic variants.

**Methods:** We performed RNA sequencing on whole blood from 3743 subjects in the COPDGene Study. RNA sequencing data from lung tissue of 1241 subjects from the Lung Tissue Research Consortium (LTRC), and whole genome sequencing data on all subjects. Associations between all SNPs within 1000 kb of a gene (cis-) and splice and gene expression quantifications were tested using tensorQTL. In COPDGene a total of 11,869,333 SNPs were tested for association with 58,318 splice clusters, and 8,792,206 SNPs were tested for association with 70,094 splice clusters in LTRC. We assessed colocalization with COPD-associated SNPs from a published GWAS[1].

**Results:** After adjustment for multiple statistical testing, we identified 28,110 splice-sites corresponding to 3,889 unique genes that were significantly associated with genotype in COPDGene whole blood, and 58,258 splice-sites corresponding to 10,307 unique genes associated with genotype in LTRC lung tissue. We found 7,576 sQTL splice-sites corresponding to 2,110 sQTL genes were shared between whole blood and lung, while 20,534 sQTL splice-sites in 3,518 genes were unique to blood and 50,682 splice-sites in 9,677 genes were unique to lung. To determine what proportion of COPD-associated SNPs were associated with transcriptional splicing, we performed colocalization analysis between COPD GWAS and sQTL data, and found that 38 genomic windows, corresponding to 38 COPD GWAS loci had evidence of colocalization between QTLs and COPD. The top five colocalizations between COPD and lung sQTLs include *NPNT*, *FBXO38*, *HHIP*, *NTN4* and *BTC*.

**Conclusions:** A total of 38 COPD GWAS loci contain evidence of sQTLs, suggesting that analysis of sQTLs in whole blood and lung tissue can provide novel insights into disease mechanisms.

## Introduction

Chronic obstructive pulmonary disease (COPD) is a complex disease characterized by irreversible airflow obstruction on lung function testing. The leading environmental risk factor for COPD is cigarette smoking, however, genetic factors have also been shown to play a role in disease susceptibility[2–5]. Genome-wide association studies (GWAS) have been used to identify genetic variants associated with COPD and lung function[1, 6–8]. However, as for most complex trait GWAS associations, the causal mechanisms are currently unknown. While it has been found that expression quantitative trait loci (eQTL) are enriched among GWAS loci, a large proportion of disease heritability remains unexplained by eQTLs [9]. Previous work has shown that splicing quantitative trait loci (sQTLs), in which genetic variants affect alternative splicing, can account for at least as many GWAS loci as eQTLs[10].

A recent genome wide association study for COPD including 35,735 cases and 222,076 controls from the UK Biobank and the International COPD Genetics Consortium identified 82 loci associated with COPD with genome wide significance[1]. Using S-PrediXcan[11], the authors discovered that 49 GWAS loci had evidence for genetically regulated expression associated with COPD using data from the Lung-eQTL consortium[1]. As S-PrediXcan is also influenced by linkage disequilibrium, most of the COPD GWAS loci are likely not explained using existing eQTLs.

Genomic loci identified as being eQTLs may also be sQTLs, as splicing is a common mechanism to alter total gene expression levels. In our previous work, we generated sQTLs in 376 subjects from the COPDGene study and found that these data could explain seven COPD GWAS associations, including the identification of *FBXO38* as a novel COPD susceptibility gene at 5q32 [10]. Here, we expand upon our findings by developing a large database of eQTLs and sQTLs in RNA from lung tissue from 1,241 subjects and in RNA from blood from 3,743 subjects followed by colocalization analysis with COPD GWAS results.

## Materials and Methods

### Study Population

This study included 3,713 non-Hispanic white and African American subjects from the COPDGene study and 1,241 subjects from the Lung Tissue Research Consortium (LTRC) (Table 1). COPDGene enrolled individuals between the ages of 45 and 80 years with a minimum of 10 pack-years of lifetime smoking history from 21 centers across the United States [12]. These subjects returned for a second study visit 5 years after the initial visit at which time they completed additional questionnaires, pre-and post-bronchodilator spirometry, chest computed tomography of the chest, and provided blood for complete blood counts (CBCs) and RNA sequencing. Samples were collected as part of the LTRC from individuals who were undergoing clinically indicated thoracic surgery procedures using a standardized protocol described in the original study design, which included pulmonary function testing, questionnaires, and chest CT [13].

**Table 1:**
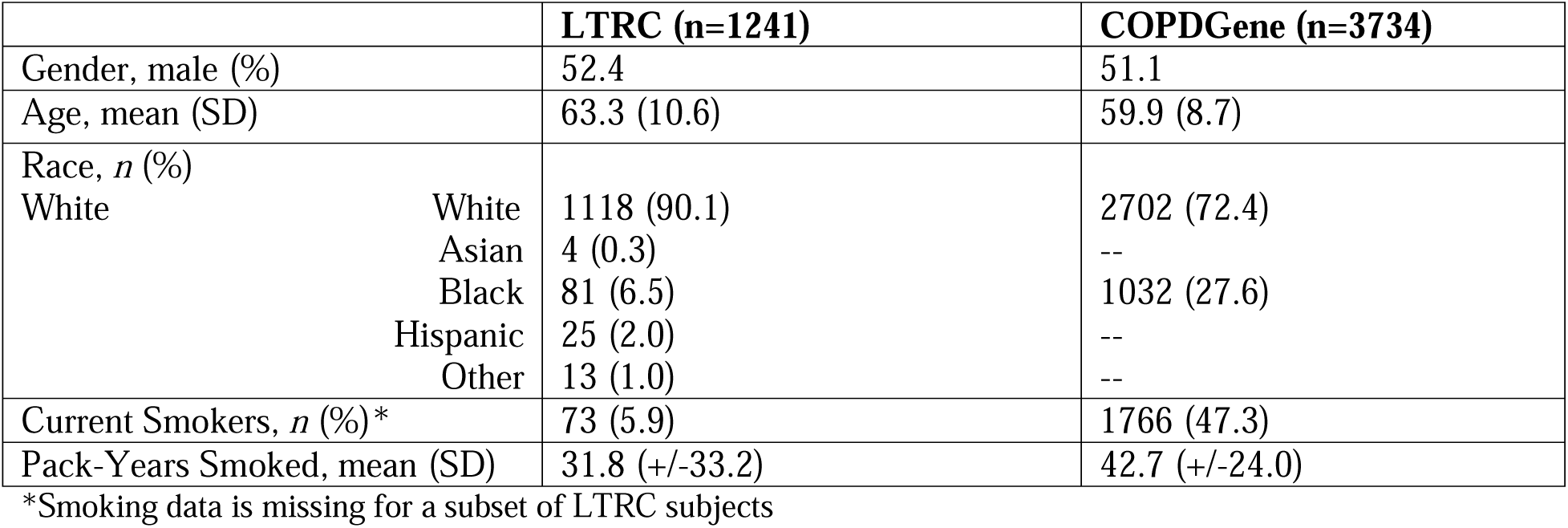
Clinical characteristics of LTRC and COPDGene study individuals included in the analysis.

### RNA sequencing, alignment and count generation

The protocols for RNASeq data generation and processing for COPDGene and LTRC have been previously described [14] [13]. Briefly, for LTRC, mRNA sequencing (RNAseq) was performed through the NHLBI TOPMed program at the University of Washington. Poly-A selection and cDNA synthesis was performed using the TruSeq Stranded mRNA kit (Illumina), and sequencing was performed on the NovaSeq6000 instrument. Sequences were aligned to GRCh38 using STAR (v2.6.1d) with the GENCODE release 29 reference. Gene-level expression quantification was performed using RSEM (v1.3.1). For COPDGene, globin reduction, ribosomal RNA depletion, and cDNA library prep was performed on total RNA from whole blood using the TruSeq Stranded Total RNA with Ribo-Zero Globin kit (Illumina, Inc., San Diego, CA), and sequencing was performed on Illumina platforms. Sequences were aligned to GRCh38 using STAR 2 pass alignment (v.2.5.2b). Gene-level expression quantification was performed using Salmon (v1.3.0) for pseudoalignment to GENCODE release 37 transcriptome, followed by summarization of isoform-level counts to gene-level counts using tximport (v.1.8.5) [15, 16]. Quantification of splicing ratios was performed in COPDGene and LTRC using Leafcutter with default parameters [17]. Intron ratios were calculated by determining how many reads support a given exon-intron junction in relation to the number of reads in that region.

### Whole Genome Sequencing

All samples were sequenced through the TOPMed program. This analysis uses Freeze 8 data.

### eQTL and sQTL Analysis

Gene expression counts were filtered to include only genes with at least 1 count per million in at least 20% of subjects, and the remaining counts were TMM normalized [18]. Leafcutter ratios were filtered to remove introns detected in less than 40% of individuals and introns with a standard deviation of less than 0.005 across subjects, and the remaining ratios were scale normalized (ie mean centered and divided by the standard error). TensorQTL [19] was used to test for association between genotypes of all SNPs within 1000 kb of the gene boundary (cis-) and quantifications of splicing or gene expression using linear models, adjusting for gender, batch, principal components of splicing or gene expression data and principal components of genetic ancestry. Calculation of principal components of genetic ancestry has been previously described [20]. A total of 8,792,206 variants (biallelic SNPs with MAF > 0.01) were tested for association with 58,258 splice sites (corresponding to 10,615 unique genes) and expression of 16,264 genes. Results were annotated using ANNOVAR [21] with annotations derived from dbSNP build 150.

### Colocalization analysis

Published GWAS data for COPD case-control status [1] were used for this analysis. Testing windows were generated by identifying all GWAS variants with p< 1x10^-5^ and calculating non-overlapping windows of 1MB on either side of each SNP. Only windows containing sQTLs or eQTLs with FDR<0.05 were tested. For each window, Bayesian colocalization tests were performed using the Moloc R package [22] to quantify the probability that the GWAS and sQTL or eQTL associations were due to a shared causal variant. Windows with a colocalization posterior probability (CPP) of greater than 0.8 were reported. Fine mapping was performed on QTL results from selected regions of interestusing SuSieR version 12.35, with in sample LD [23].

### Long read RNA-seq analysis in human lung samples from the LTRC

We conducted targeted Oxford Nanopore Technologies (ONT) long read sequencing on RNA from 170 human lung samples from the LTRC on genes selected from colocalization analysis. The enrichment and library generation procedures are described in detail in the Supplemental Methods. The final library was loaded onto a PromethION R10.4 flow cell and run at 400bp/s. Approximately once per day, flow cells were flushed and treated with DNAse I, then loaded with additional library to increase sequencing throughput. Resulting raw reads were basecalled using the SUP model of guppy (v6) and consensus called and demultiplexed using C3POa (v2.3). R2C2 reads were analyzed to identify and quantify isoforms using version 3.5 of Mandalorion (https://github.com/rvolden/Mandalorion-Episode-III). Mandalorian was run twice to identify high abundance isoforms (>10% of total isoform expression; -O 0,40,0,40 -r 0.1 -i 1 - w 1 -n 2 -R 5) and high and low abundance isoforms (-O 0,40,0,40 -r 0.01 -i 1 -w 1 -n 2 -R 5).

## Results

### Quantification of gene expression and RNA splicing in COPDGene blood and LTRC lung

Using RNASeq data from LTRC lung tissue (n=1241) and COPDGene whole blood (n=3713) we identified splice sites using Leafcutter [24], which identifies and quantifies intron exclusion. After filtering out splice sites with low usage, we identified a total of 223,128 splice sites, corresponding to 15,121 genes in LTRC, and 160,658 splice sites corresponding to 12,096 genes in COPDGene (Table 2). In both LTRC and COPDGene, the majority (50.1% and 51.0%, respectively) of identified splice sites were annotated in GENCODE, followed by cryptic 3’, cryptic 5’, cryptic unanchored splice sites (meaning both splice donor and acceptor were unannotated) and novel annotated pairs (Table 3). Gene expression of 16,266 genes in LTRC and 15,507 genes in COPDGene met expression thresholds (Table 2).

**Table 2:**
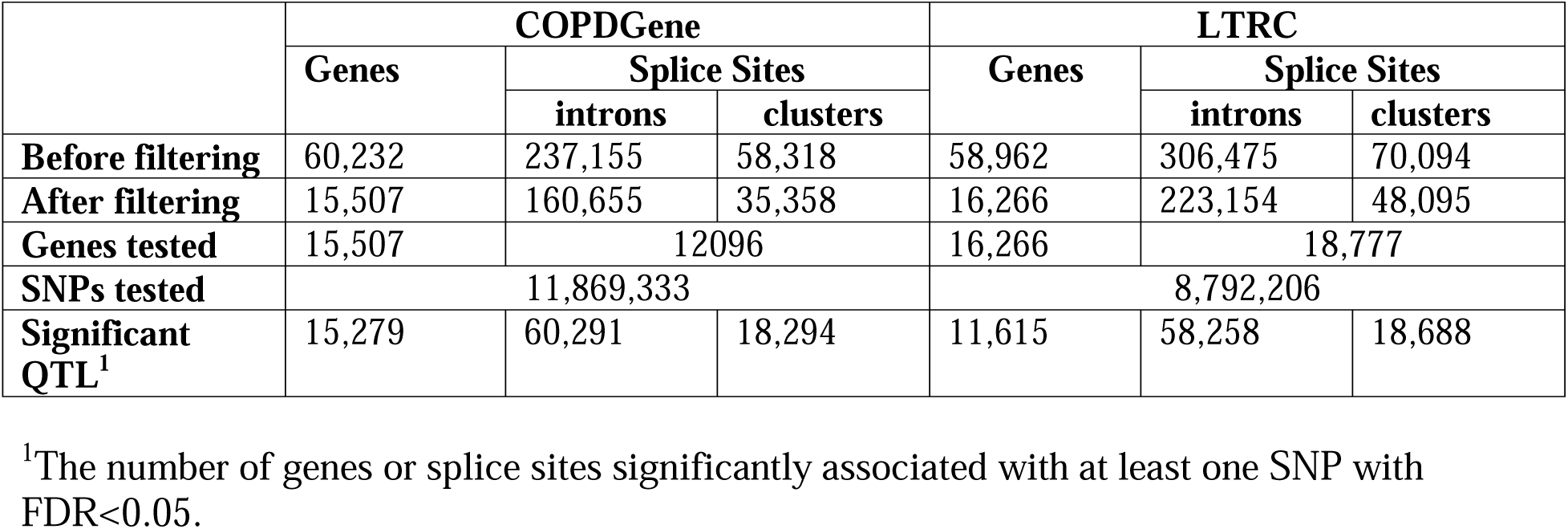
Summary of expressionQTLs and spliceQTLs tested.

|  | COPDGene |  |  | LTRC |  |  |
| --- | --- | --- | --- | --- | --- | --- |
|  | Genes | Splice Sites |  | Genes | Splice Sites |  |
|  |  | introns | clusters |  | introns | clusters |
| Before filtering | 60,232 | 237,155 | 58,318 | 58,962 | 306,475 | 70,094 |
| After filtering | 15,507 | 160,655 | 35,358 | 16,266 | 223,154 | 48,095 |
| Genes tested | 15,507 | 12096 |  | 16,266 | 18,777 |  |
| SNPs tested | 11,869,333 |  |  | 8,792,206 |  |  |
| Significant QTL<sup>1</sup> | 15,279 | 60,291 | 18,294 | 11,615 | 58,258 | 18,688 |

<sup>1</sup>The number of genes or splice sites significantly associated with at least one SNP with FDR<0.05.

**Table 3:** Annotations of Leafcutter Splice Sites Identified in COPDGene and LTRC.

|  | **Frequency in LTRC** | **Percent in LTRC** | **Frequency in COPDGene** | **Percent in COPDGene** |
| --- | --- | --- | --- | --- |
| **Annotated<sup>1</sup>** | 113562 | 50.9 | 81111 | 50.5 |
| **Cryptic 5'** | 30166 | 13.5 | 21232 | 13.2 |
| **Cryptic 3'** | 32258 | 14.5 | 24047 | 15.0 |
| **Cryptic unanchored** | 8504 | 3.8 | 5788 | 3.6 |
| **Novel annotated pair** | 17991 | 8.1 | 13293 | 8.3 |
| **Unknown strand** | 20626 | 9.2 | 15166 | 9.4 |

<sup>1</sup>Annotated = both 5' and 3' splice sites have been previously annotated; Cryptic fiveprime = the 5' splice site is not annotated but the 3' splice site is annotated; Cryptic threeprime = the 3' splice site is not annotated but the 5' splice site is annotated; Cryptic unanchored = neither splice site has been previously annotated; Novel annotated pair = both 5' and 3' splice sites have been individually annotated but the combination have not been annotated as a junction; Unknown strand = it is not possible to determine the directionality of the splice sites based on the RNASeq read.

### Identification of eQTL and sQTLs in human lung tissue and whole blood

We next tested for association between genotype and gene expression or splicing to identify eQTLs and sQTLs in lung and blood. In LTRC lung tissue we identified 58,258 splice sites (corresponding to 10,615 genes) associated with at least one SNP with q-value<0.05; in COPDGene blood 60,291 splice sites (corresponding to 8,671 genes) were associated with at least one SNP (Table 2). In addition, we identified 12,225 genes associated with at least one SNP (eQTLs) in LTRC, and 15,279 eQTL genes in COPDGene. We found that 14,064 sQTL splice-site-SNP pairs (13%) were found in both blood and lung, while 6,353 eQTL gene-SNP pairs were shared between both tissues (25%) (Figure 1). In addition, we found that 5,787 sQTL gene-SNP pairs overlapped with eQTL gene-SNP pairs in LTRC (47%), while 7,455 sQTL gene-SNP pairs overlapped with eQTL gene-SNP pairs in COPDGene (49%).

**Figure 1:**
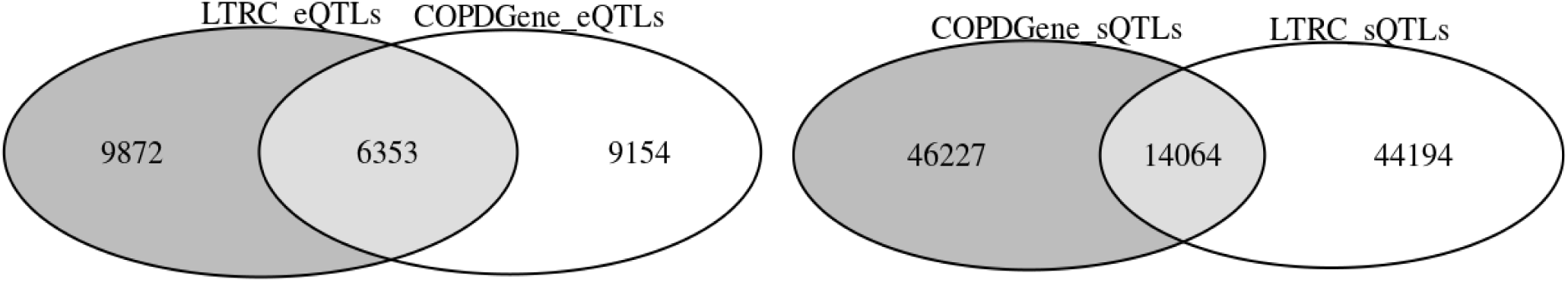
Overlap between LTRC lung and COPDGene blood eQTLs and sQTLs that were significant at FDR<0.05

### Functional annotation of sQTLs and eQTLs

Next we categorized eQTLs and sQTLs-SNPs on the basis of their location relative to the gene with which they were associated (Table 4). The genomic distribution of eQTLs and sQTLs was similar, with the largest proportion of variants located upstream of the gene region in both COPDGene (32.1% and 34.7 %, respectively) and LTRC (31.8% and 29.4%, respectively). The next most frequent SNPs were intronic, downstream of gene, intergenic, and 3’ UTR variants. Only a small percentage of lead sQTL-SNPs directly modified a splice site. There was no difference in variant position in either eQTL vs sQTL and in COPDGene blood vs LTRC lung.

**Table 4:**
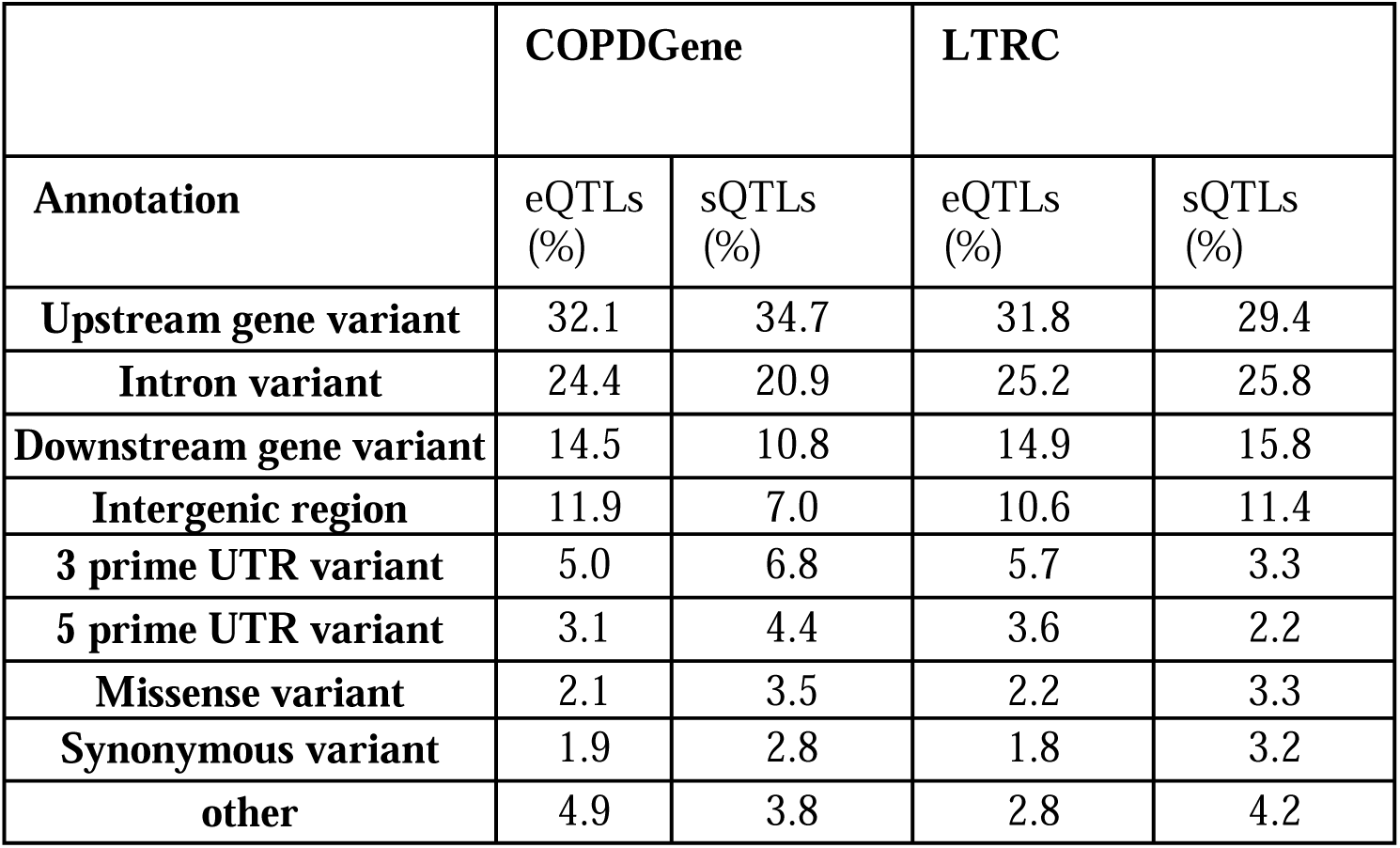
Annotation of QTL variants in relation to the gene body.

|  | **COPDGene** |  | **LTRC** |  |
| --- | --- | --- | --- | --- |
| **Annotation** | eQTLs (%) | sQTLs (%) | eQTLs (%) | sQTLs (%) |
| **Upstream gene variant** | 32.1 | 34.7 | 31.8 | 29.4 |
| **Intron variant** | 24.4 | 20.9 | 25.2 | 25.8 |
| **Downstream gene variant** | 14.5 | 10.8 | 14.9 | 15.8 |
| **Intergenic region** | 11.9 | 7.0 | 10.6 | 11.4 |
| **3 prime UTR variant** | 5.0 | 6.8 | 5.7 | 3.3 |
| **5 prime UTR variant** | 3.1 | 4.4 | 3.6 | 2.2 |
| **Missense variant** | 2.1 | 3.5 | 2.2 | 3.3 |
| **Synonymous variant** | 1.9 | 2.8 | 1.8 | 3.2 |
| **other** | 4.9 | 3.8 | 2.8 | 4.2 |

### Colocalization of QTLs with COPD Case-Control GWAS data

We next sought to identify eQTLs and sQTLs which contribute to COPD risk by performing genetic colocalization between the QTL data and COPD case-control GWAS data. Based on GWAS data, 239 genomic windows with COPD GWAS p-values < 5x10^-5^ were identified, and of these, 237 contained sQTLs or eQTLs with q-value<0.05. We also included colocalization results for 3’ UTR alternative polyadenylation QTLs (apaQTLs) from our previous study [25]. We identified 38 windows with a colocalization posterior probability of association (PPA) > 0.8 with either eQTL, sQTL or apaQTL data and GWAS p-value < 5.0x10^-8^ (Table 5). We found that for 19 loci the strongest colocalization (largest PPA) with GWAS data was in LTRC, and for 19 loci the strongest colocalization was in COPDGene. In LTRC, the largest number of GWAS loci colocalized with sQTLs (9 loci), followed by eQTLs (7 loci) then apaQTLs (3 loci). In COPDGene, the majority of colocalizations were with sQTLs (15 loci), with only one 1 locus colocalizing most strongly with eQTLs, and 3 with apaQTLs. We compared the colocalization findings with the target genes identified from the original GWAS analysis to determine whether the QTLs identified novel targets from previous analyses. We found that for 7 loci all genes identified in the current QTL analysis were previously identified, and for 26 loci one or more new targets were found (Supplementary Table 1). For further characterization we focused on sQTL colocalizations in LTRC lung, with the top five colocalizations (by highest PPA) being *Nephronectin* (*NPNT)*, *F-box protein 38* (*FBXO38)*, *Hedgehog interacting protein* (*HHIP)*, *Netrin 4 (NTN4)* and *Betacellulin* (*BTC)*. We have previously published on NPNT colocalizations in the lung [26], and for HHIP significant evidence suggests that the mechanism underlying the association is an eQTL effect [27]. *NTN4* appears to be a promoter usage eQTL instead of an sQTL. Therefore, we highlight the results for *FBXO38* and *BTC* below. All colocalization results are available online at (https://copd-moloc.bwh.harvard.edu/).

**Table 5:** Summary of Colocalization Analysis.

| Best Colocalized SNP<sup>1</sup> | PPA<sup>2</sup> | Lead QTL |  |  |  |  | GWAS |  |
| --- | --- | --- | --- | --- | --- | --- | --- | --- |
|  |  | Gene<sup>2</sup> | QTL Type | Splice site | p-value | Effect<sup>3</sup> | p-value | Effect<sup>3</sup> |
| 4:144567946:A:G | 0.973 | *HHIP* | LTRC_sQTL | 4:144734889-144737129 | 0.00610 | 0.104 | 4.09E-59 | 16.136 |
| 4:105897896:G:A | 0.999 | *NPNT* | LTRC_sQTL | 4:105927428-105931515 | 8.12E-10 | 0.114 | 3.04E-46 | 14.331 |
| 5:148475407:C:T | 0.973 | *FBXO38* | LTRC_sQTL | 5:148414306-148415928 | 0.00333 | -0.195 | 2.58E-33 | -12.010 |
| 15:71329185:G:A | 0.992 | *THSD4* | LTRC_eQTL | ENSG00000187720.14 | 0.154 | 4.653 | 1.58E-32 | 11.852 |
| 16:75439564:G:A | 0.933 | *TMEM170A* | COPDGene_sQTL | 16:75451839-75464232 | 7.63E-08 | 0.187 | 1.26E-20 | 9.277 |
| 6:30814428:G:C | 0.987 | *IER3* | COPDGene_sQTL | 6:30744196-30744279 | 0.000119 | -0.176 | 1.41E-20 | 9.295 |
| 4:88948181:T:C | 0.832 | *PKD2* | COPDGene_sQTL | 4:88019571-88035071 | 3.76E-08 | -0.518 | 4.23E-18 | -8.722 |
| 6:32660630:T:C | 0.997 | *HLA-DRB5* | COPDGene_sQTL | 6:32520345-32555661 | 6.09E-15 | -0.275 | 5.56E-18 | -8.661 |
| 5:157505976:A:T | 0.968 | *ADAM19* | LTRC_eQTL | ENSG00000135074.15 | 8.62E-10 | 2.468 | 1.22E-16 | -8.324 |
| 3:128242335:T:A | 0.935 | *EEFSEC* | COPDGene_sQTL | 3:128195329-128246836 | 0.0014 | 0.032 | 3.53E-15 | 7.897 |
| 6:29639324:T:C | 0.997 | *HLA-E* | COPDGene_sQTL | 6:30490515-30491137 | 4.82E-301 | -0.890 | 4.35E-15 | -7.864 |
| 6:27420975:T:C | 0.976 | *ZKSCAN4* | COPDGene_sQTL | 6:28249834-28251867 | 5.35E-20 | 0.308 | 3.58E-14 | -7.573 |
| 6:28651576:A:G | 0.996 | *GABBR1* | COPDGene_eQTL | ENSG00000204681.11 | 3.96E-17 | -2.034 | 3.82E-14 | -7.556 |
| 6:26409662:G:C | 0.905 | *BTN2A1* | LTRC_sQTL | 6:26459828-26463244 | 1.79E-10 | -0.486 | 1.03E-13 | 7.449 |
| 6:32775967:G:A | 0.982 | *HLA-DRB5* | COPDGene_sQTL | 6:32481801-32525584 | 8.02E-09 | -0.222 | 3.76E-12 | 6.955 |
| 2:238965524:G:A | 0.941 | *TWIST2* | LTRC_eQTL | ENSG00000233608.4 | 0.0483 | 0.269 | 2.17E-11 | -6.699 |
| 3:25496178:G:A | 0.990 | *RARB* | LTRC_eQTL | ENSG00000077092.19 | 2.60E-06 | 0.925 | 5.84E-11 | -6.570 |
| 2:9145396:G:A | 0.956 | *LINC00299* | COPDGene_sQTL | 2:8299793-8312726 | 0.0479 | -0.115 | 2.41E-10 | 6.343 |
| 1:16979534:C:A | 0.990 | *CROCC* | COPDGene_sQTL | 1:16922798-16924325 | 0.000528 | -0.121 | 6.55E-10 | 6.192 |
| 7:100032719:C:T | 0.994 | *ZSCAN21* | COPDGene_sQTL | 7:100049841-100051478 | 6.28E-05 | -0.153 | 7.26E-10 | -6.175 |
| 19:45790878:G:A | 0.998 | *DMWD* | LTRC_APAQTL | ENST00000597053.1 | 0.00118 | -0.044 | 1.67E-09 | -6.009 |
| 1:39582337:G:A | 0.933 | *PPIEL* | COPDGene_sQTL | 1:39548951-39554349 | 0.0176 | -0.128 | 1.98E-09 | 6.009 |
| 12:95843792:T:C | 0.972 | *NTN4* | LTRC_sQTL | 12:95713338-95717545 | 6.56E-09 | -0.400 | 2.64E-09 | -5.935 |
| 4:74748514:C:T | 0.951 | *BTC* | LTRC_sQTL | 4:74748149-74750573 | 0.00187 | -0.238 | 3.45E-09 | -5.903 |
| 1:111195294:T:C | 0.850 | *DENND2D* | COPDGene_sQTL | 1:111194726-111195668 | 0.00843 | -0.556 | 3.59E-09 | -5.892 |
| 14:92649065:G:C | 0.963 | *RIN3* | LTRC_APAQTL | ENST00000553992.1 | 0.00115 | -0.025 | 6.69E-09 | 5.795 |
| 1:239689643:G:C | 0.968 | *CHRM3-AS2* | COPDGene_APAQTL | ENST00000593855 | 2.77E-48 | -0.279 | 9.61E-09 | 5.755 |
| 11:13145018:T:C | 0.941 | *RASSF10* | LTRC_eQTL | ENSG00000189431.7 | 4.27E-09 | 0.250 | 1.18E-08 | 5.717 |
| 5:151215512:A:G | 0.994 | *GM2A* | LTRC_APAQTL | ENST00000523004.1 | 9.43E-100 | 0.011 | 1.40E-08 | -5.695 |
| 7:2830820:T:G | 0.873 | *GNA12* | LTRC_sQTL | 7:2733501-2762829 | 0.00185 | 0.156 | 1.74E-08 | 5.648 |
| 17:38730575:G:A | 0.990 | *CISD3* | LTRC_eQTL | ENSG00000277972.1 | 8.70E-11 | -2.872 | 1.90E-08 | 5.620 |
| 3:29430921:G:C | 0.985 | *RBMS3* | LTRC_eQTL | ENSG00000144642.21 | 1.02E-11 | -2.128 | 2.04E-08 | 5.636 |
| 15:49578340:A:G | 0.943 | *FAM227B* | LTRC_sQTL | 15:49589745-49606156 | 0.0146 | -0.151 | 2.82E-08 | 5.564 |
| 16:58063696:G:C | 0.890 | *MMP15* | LTRC_sQTL | 16:58040698-58041617 | 0.0122 | 0.274 | 4.26E-08 | 5.500 |
| 10:80458350:G:C | 0.995 | *TSPAN14* | COPDGene_sQTL | 10:80459507-80463088 | 3.64E-07 | 0.132 | 4.30E-08 | -5.465 |
| 1:45543641:G:C | 0.959 | *TESK2* | COPDGene_APAQTL | ENST00000486676 | 0.00967 | -0.032 | 4.67E-08 | -5.485 |
| 2:42206107:C:T | 0.980 | *COX7A2L* | COPDGene_APAQTL | ENST00000463055 | NA | NA | 4.85E-08 | -5.446 |
| 17:30129740:C:T | 0.865 | *NSRP1* | COPDGene_sQTL | 17:30118173-30156218 | 0.000150 | 0.116 | 4.86E-08 | 5.480 |

<sup>1</sup>SNP = chromosome, position, reference, and effect allele, where position is build 38.
<sup>2</sup>PPA = posterior probability of association from moloc software. For some loci, there were multiple colocalizations between QTLs and GWAS data, the results for the association with the highest PPA is reported here
<sup>3</sup>The direction of the effect size is based on the second allele listed in the variant ID

### Characterization of sQTL for FBXO38

We first sought to replicate the association we previously characterized in 365 subjects from COPDGene between the rs7730971 (5:148411297:C:G) variant and *FBXO38* splicing in whole blood which colocalized with COPD GWAS findings [10]. Here, we identified two splicing clusters (ie a group of splice junctions with shared start or stop positions [24]) in lung tissue in *FBXO38* which were associated with COPD-related variants. First, we confirmed our previous finding that rs7730971 was significantly associated with inclusion of a 158 bp cryptic exon located between exons 9 and 10 (Figure 2). While significant colocalization was not detected using moloc, we previously found colocalization in this locus using eCaviar [28]. The G allele, which is associated with increased risk of COPD, is also associated with increased inclusion of the cryptic exon (Figure 2b). SpliceAI [29] indicates that rs7730971-G is predicted to slightly increase the splice acceptor strength of a splice site 217 bp upstream of the variant, which corresponds to the 5’ splice acceptor of the cryptic exon, confirming that this is the likely causal variant. Using long-read sequencing we identified one isoform meeting expression thresholds which includes the 158 bp cryptic exon (Figure 2c). This transcript includes a premature stop codon in the cryptic exon, and because this stop is more than 50 bases from the transcriptional stop, it would likely be subject to nonsense mediated decay [30]. This isoform is more abundant in the GG genotype compared to AA (5 reads vs 1). Supporting the finding that the transcript containing the cryptic exon is subject to nonsense mediated decay, we found that rs7730971 is also an eQTL for *FBXO38*, with the G allele associated with decreased expression (Figure 2d). We additionally identified a new colocalization between genetic variants in the *FBXO38/HTR4* region and inclusion of an exon at chr5: 148415241-148415387 (Supplementary Data).

**Figure 2:**
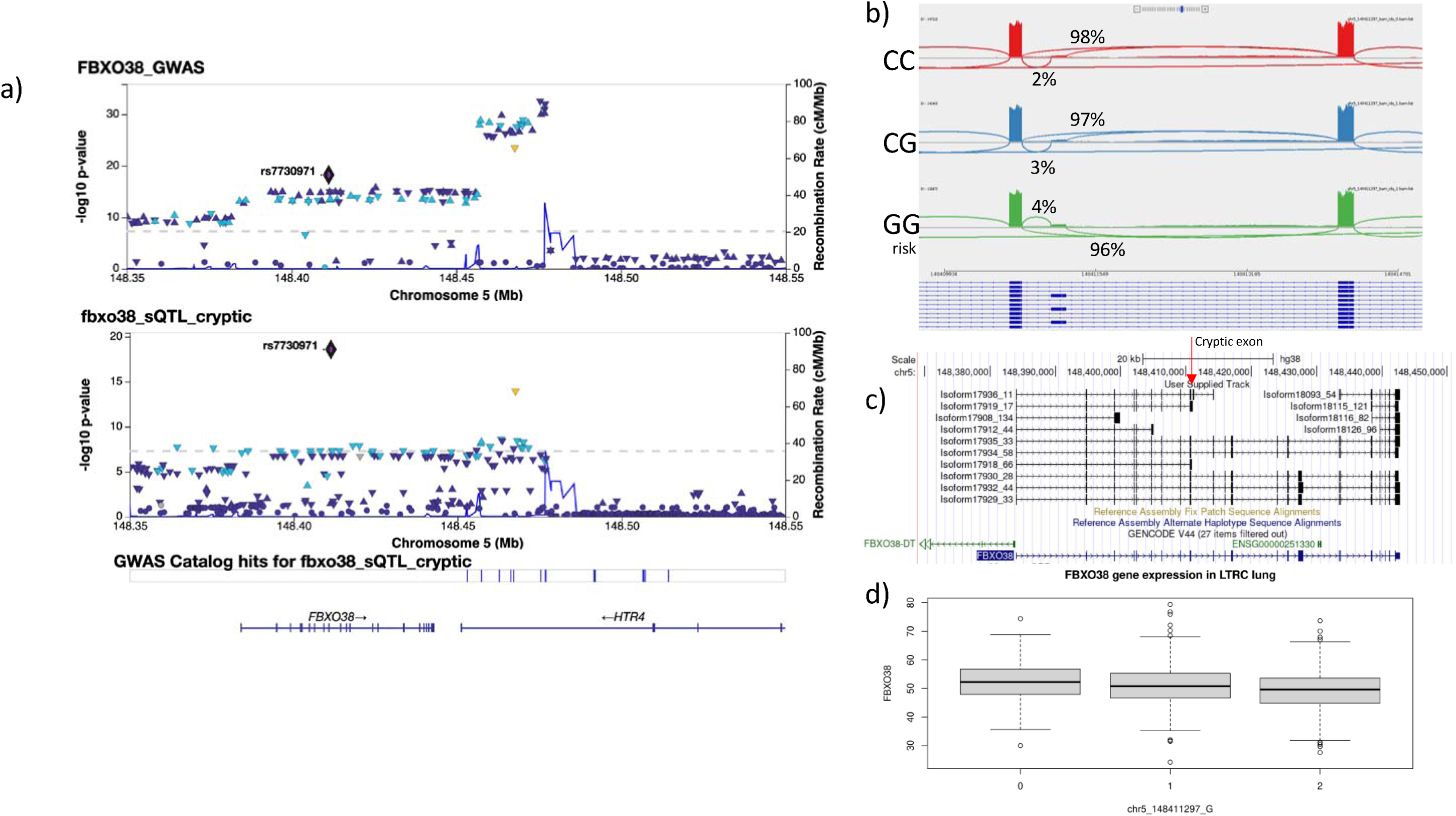
Replication of *FBXO38* sQTL findings in lung tissue. a) Locus association plot for COPD GWAS and *FBXO38* sQTL. The lead SNP associated with FBXO38 splicing, rs7730971, is highlighted in purple and used as the LD reference. b) IGV sashimi plot showing the region spanning chr5:148409934-148414781 for 191 subjects from each genotype of rs7730971. c) Long read sequencing data showing all FBXO38 isoforms representing at least 1% of total FBXO38 expression. The cryptic exon is identified with a red arrow. D) Boxplot of total gene expression values for *FBXO38*.

### Colocalization analysis of a genetic signal at BTC

Another significant colocalization between sQTLs and COPD GWAS findings was identified in LTRC lung tissue at the *BTC* locus where we found evidence of a shared genetic signal between variants associated with alternative inclusion of exon 4 of *BTC*, and COPD risk GWAS data (PPA=0.95) (Figure 3a). The lead colocalized variant was rs62316278 (4:74748514:C:T), and the COPD risk allele (C) is associated with increased inclusion of exon 4 (Figure 3b). This variant is within the 95% credible set for sQTL data using SuSie fine mapping, along with 42 additional variants. Of the variants in the 95% credible set, we found using SpliceAI that rs11938093-T (4:74750631:A:T) is associated with the gain of a splice donor 58 bp upstream of the SNP, corresponding to the splice donor of exon 4, as well as the gain of a splice acceptor 88 bp downstream of the SNP, corresponding to the splice acceptor of exon 4. Long read sequencing identified four high abundance isoforms for *BTC* (representing at least 10% of BTC expression each) (Figure 3c), and the proportion of isoforms containing exon 4 was higher in CC vs TT subjects (84% vs 48%, p=0.0003). These isoforms correspond to NM_001729 (BTC-201, ENST00000395743.8) and NM_001316963 (not included in ENSEMBL). NM_001729 (including exon 4) is the primary version of BTC, and codes for a 178 amino acid protein, while NM_001316963 codes for 129 amino acids.

**Figure 3:**
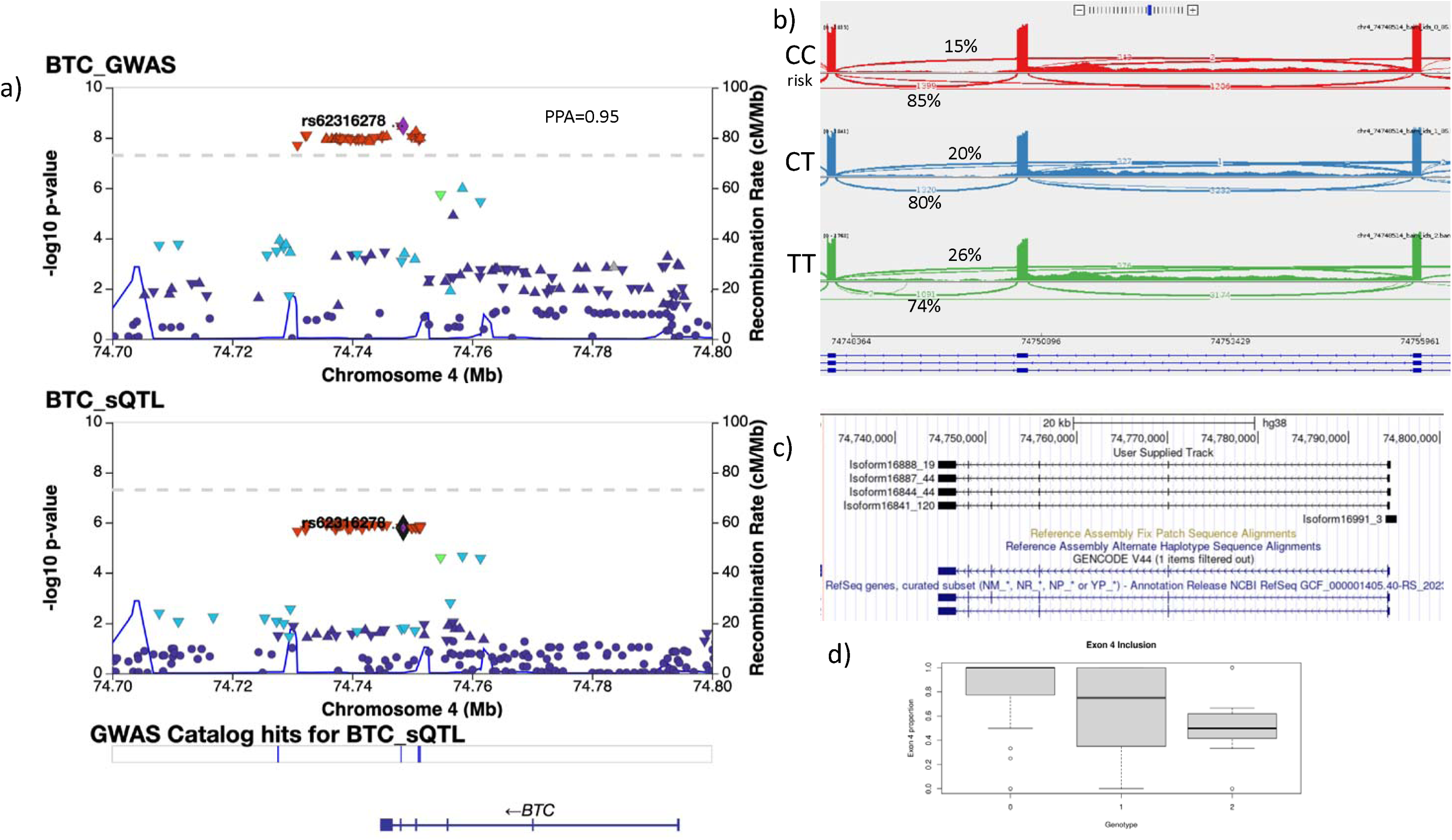
sQTLs for *BTC* colocalize with GWAS data for COPD. a) Locus association plot for COPD GWAS and *BTC* sQTL. The lead colocalized SNP, rs62316278, is highlighted in purple and used as the LD reference. b) IGV sashimi plot showing the region spanning chr4:74748364-74755961 for 86 subjects from each genotype of rs62316278. c) Long read sequencing data showing all BTC isoforms representing at least 10% of total BTC expression. D) Proportion of BTC expression of isoforms containing exon 4 by copies of rs62316278-T.

## Discussion

In this study we build upon our previous work characterizing COPD associated sQTLs in blood RNASeq data from COPDGene by generating a large dataset of eQTLs and sQTLs in human blood and lung tissue and identifying gene expression and splicing events, and we identify a substantial number of QTLs that suggest a functional mechanism for COPD GWAS loci. We found that approximately 50% of splice sites identified were not currently annotated, indicating the vast amount of currently uncharacterized splicing variability present in the transcriptome.

Among the 223,128 splice sites identified in LTRC lung tissue and 160,658 in COPDGene blood, we found that 58,258 (26%) and 60,291 (38%) splice sites, respectively, were associated with at least one variant within 1 MB (FDR<5%). In addition, of the 16,266 genes expressed in LTRC and 15,507 genes expressed in COPDGene, 12,225 (75%) and 15,279 (99%) of genes, respectively, were associated with at least one SNP. The majority of eQTL and sQTL SNPs were located upstream of the gene body or in intronic regions, suggesting that many sQTLs function through long range or indirect effects, as opposed to modifying splice donors or acceptors directly. We identified 38 loci (corresponding to 33 of the original 82 GWAS loci) with significant colocalization with either eQTL, sQTL or apaQTL data, and of these, 9 loci colocalized most strongly with LTRC sQTLs, and 15 with COPDGene sQTLs. We confirmed our previous sQTL findings in the *FBXO38/HTR4* region, and identified *BTC* as additional novel target with strong COPD colocalization.

Here, we validated in lung tissue our previous findings from blood that rs7730971 is associated with splicing of a cryptic exon in *FBXO38*. Open reading frame analysis of the full length transcript sequence indicates that the cryptic exon contains a premature stop codon, and therefore this transcript is likely subject to nonsense mediated decay. We also found that rs7730971 is associated with gene expression of *FBXO38*, with decreased expression with the G allele, which is also the variant associated with a bioinformatically predicted increase in nonsense mediated decay. Therefore, the likely mechanism underlying the eQTL association is degradation of *FBXO38* as result of increased inclusion of a cryptic exon which results in a transcript with an early stop codon. This is an example of a mechanism by which an eQTL can be mediated through an sQTL.

The allele associated with decreased expression of *FBXO38* is associated with increased COPD risk, suggesting that *FBXO38* plays a protective role against COPD. We found an additional colocalization between rs10037493, the most significant COPD GWAS SNP in the locus, and an additional *FBXO38* exon. The long read sequences containing this exon correspond to two predicted isoforms, each encoding shorter proteins than the most abundant isoform. The COPD risk allele is associated with the shortened isoforms, indicating that the full length *FBXO38* isoform is protective. These shortened isoforms lack the F-box domain, which is a component of all members of F-box proteins, including FBXO38. F-box proteins are a component of the ubiquitin ligase complex and also function as transcription factors [31].

FBXO38 specifically is a coactivator of the Kruppel-like factor 7 (KLF7) zinc finger transcription factor [32, 33]. While little is known about the function of FBXO38 and its potential role in COPD, the lack of the F-box protein, which is responsible for protein-protein interactions, in disease associated isoforms indicates that FBXO38 interactions are critical for protection against COPD.

We identified an additional colocalization between the COPD GWAS and a novel candidate gene, *BTC*. We specifically found that rs62316278, the lead GWAS SNP in the *BTC* region, is also associated with splicing of exon 4, with the risk allele (C), increasing exon 4 inclusion. The primary form of *BTC*, NM_001729, includes exon 4, and rs62316278-C is associated with an increased proportion of NM_001729 relative to NM_001316963, a protein coding isoform which lacks exon 4. This suggests that reduced inclusion of exon 4 is protective for COPD. BTC is a member of the epidermal growth factor (EGF) family of peptide ligands, and a ligand for epidermal growth factor receptor (EGFR). Human *BTC* encodes a 178-amino acid product corresponding to the BTC precursor protein (pro-BTC) and contains several domains including a signal peptide, an EGF motif and transmembrane domains [34]. The mature sequence of BTC is cleaved from the extracellular domain of BTC to produce an 80 amino acid protein. Based on the structure of other members of the EGF family, exon 4 is predicted to make up the third loop of the EGF-like motif and the transmembrane domain [34]. The EGF domain is critical to binding with EGF ligands, and therefore the isoform lacking exon 4 could be predicted to have impaired function. Several previous studies have linked *BTC* expression to COPD, and *BTC* has been found to be higher in ex-smokers with COPD than without COPD, has been associated with emphysema in alpha-1 antitrypsin deficiency [35] and has been found elevated in acute exacerbations in COPD [36]. More targeted work is needed to investigate the function of *BTC* isoforms in COPD.

The major strength of this study is the large sample size which allowed us to comprehensively characterize alternative splicing in whole blood and lung tissue. Our sample size significantly exceeds that of other commonly used resources such as The Genotype Tissue Expression Project (GTEx), which includes 515 lung samples and 670 blood samples. While we sought colocalization with GOLD-defined COPD, we expect that other respiratory- or smoking-related genetic associations would benefit from the use of this resource. One potential weakness is that our RNASeq was performed using whole lung tissue and whole blood samples, which contain a variety of cell types. Therefore, some of the changes in transcriptional splicing detected may actually be reflecting differences in inter-individual cell proportions. Another limitation is the use of Moloc, which attempts colocalization with the most significant signal in the region, and may not be optimal in the setting of multiple QTL or GWAS signals in the region. Additional work characterizing splicing using single-cell short or long read RNASeq is required.

In conclusion, we discovered that multiple COPD GWAS associations colocalize with sQTLs, and identify or replicate several candidate genes as COPD targets for follow-up.

## Data Availability

LTRC genotyping data are available on dbGaP with accession number phs001662.v1.p1
LTRC RNA-seq data are available through TOPMed, https://topmed.nhlbi.nih.gov.COPDGene data are available on dbGaP with accession numbers phs000179 and phs000765.

https://copd-moloc.bwh.harvard.edu/

## Funding/Acknowledgements

This work was funded by K01HL157613, R01HL157879, P01HL114501, X01HL139404, R01HL124233, R01HL126596, R01HL153248, R01HL149861, R01 HL111527HL135142, and NIGMS R35 GM140844.

Molecular data for the Trans-Omics in Precision Medicine (TOPMed) program was supported by the National Heart, Lung and Blood Institute (NHLBI). Whole Genome Sequencing and RNASeq for "NHLBI TOPMed: The Lung Tissue Research Consortium (phs001662)” was performed at Northwest Genome Center (NWGC, HHSN268201600032I, RNASeq) and Broad Genomics (HHSN268201600034I, WGS) Core support including centralized genomic read mapping and genotype calling, along with variant quality metrics and filtering were provided by the TOPMed Informatics Research Center (3R01HL-117626-02S1; contract HHSN268201800002I). Core support including phenotype harmonization, data management, sample-identity QC, and general program coordination were provided by the TOPMed Data Coordinating Center (R01HL-120393; U01HL-120393; contract HHSN268201800001I). We gratefully acknowledge the studies and participants who provided biological samples and data for TOPMed..

The COPDGene study (NCT00608764) is supported by grants from the NHLBI (U01HL089897 and U01HL089856), by NIH contract 75N92023D00011, and by the COPD Foundation through contributions made to an Industry Advisory Committee that has included AstraZeneca, Bayer Pharmaceuticals, Boehringer-Ingelheim, Genentech, GlaxoSmithKline, Novartis, Pfizer and Sunovion.

This study utilized biological specimens and data provided by the Lung Tissue Research Consortium (LTRC) supported by the National Heart, Lung, and Blood Institute (NHLBI).

## COPDGene® Investigators – Core Units

*Administrative Center*: James D. Crapo, MD (PI); Edwin K. Silverman, MD, PhD (PI); Barry J. Make, MD; Elizabeth A. Regan, MD, PhD

*Genetic Analysis Center*: Terri H. Beaty, PhD; Peter J. Castaldi, MD, MSc; Michael H. Cho, MD, MPH; Dawn L. DeMeo, MD, MPH; Adel El Boueiz, MD, MMSc; Marilyn G. Foreman, MD, MS; Auyon Ghosh, MD; Lystra P. Hayden, MD, MMSc; Craig P. Hersh, MD, MPH; Jacqueline Hetmanski, MS; Brian D. Hobbs, MD, MMSc; John E. Hokanson, MPH, PhD; Wonji Kim, PhD; Nan Laird, PhD; Christoph Lange, PhD; Sharon M. Lutz, PhD; Merry-Lynn McDonald, PhD; Dmitry Prokopenko, PhD; Matthew Moll, MD, MPH; Jarrett Morrow, PhD; Dandi Qiao, PhD; Elizabeth A. Regan, MD, PhD; Aabida Saferali, PhD; Phuwanat Sakornsakolpat, MD; Edwin K. Silverman, MD, PhD; Emily S. Wan, MD; Jeong Yun, MD, MPH

*Imaging Center*: Juan Pablo Centeno; Jean-Paul Charbonnier, PhD; Harvey O. Coxson, PhD; Craig J. Galban, PhD; MeiLan K. Han, MD, MS; Eric A. Hoffman, Stephen Humphries, PhD; Francine L. Jacobson, MD, MPH; Philip F. Judy, PhD; Ella A. Kazerooni, MD; Alex Kluiber; David A. Lynch, MB; Pietro Nardelli, PhD; John D. Newell, Jr., MD; Aleena Notary; Andrea Oh, MD; Elizabeth A. Regan, MD, PhD; James C. Ross, PhD; Raul San Jose Estepar, PhD; Joyce Schroeder, MD; Jered Sieren; Berend C. Stoel, PhD; Juerg Tschirren, PhD; Edwin Van Beek, MD, PhD; Bram van Ginneken, PhD; Eva van Rikxoort, PhD; Gonzalo Vegas SanchezFerrero, PhD; Lucas Veitel; George R. Washko, MD; Carla G. Wilson, MS;

*PFT QA Center*, Salt Lake City, UT: Robert Jensen, PhD

*Data Coordinating Center and Biostatistics, National Jewish Health, Denver, CO*: Douglas Everett, PhD; Jim Crooks, PhD; Katherine Pratte, PhD; Matt Strand, PhD; Carla G. Wilson, MS Epidemiology Core, University of Colorado Anschutz Medical Campus, Aurora, CO: John E. Hokanson, MPH, PhD; Erin Austin, PhD; Gregory Kinney, MPH, PhD; Sharon M. Lutz, PhD; Kendra A. Young, PhD Version Date: March 26, 2021

*Mortality Adjudication Core*: Surya P. Bhatt, MD; Jessica Bon, MD; Alejandro A. Diaz, MD, MPH; MeiLan K. Han, MD, MS; Barry Make, MD; Susan Murray, ScD; Elizabeth Regan, MD; Xavier Soler, MD; Carla G. Wilson, MS

*Biomarker Core*: Russell P. Bowler, MD, PhD; Katerina Kechris, PhD; Farnoush BanaeiKashani, PhD

## COPDGene® Investigators – Clinical Centers

*Ann Arbor VA*: Jeffrey L. Curtis, MD; Perry G. Pernicano, MD

*Baylor College of Medicine, Houston, TX*: Nicola Hanania, MD, MS; Mustafa Atik, MD; Aladin Boriek, PhD; Kalpatha Guntupalli, MD; Elizabeth Guy, MD; Amit Parulekar, MD;

*Brigham and Women’s Hospital, Boston, MA*: Dawn L. DeMeo, MD, MPH; Craig Hersh, MD, MPH; Francine L. Jacobson, MD, MPH; George Washko, MD

*Columbia University, New York, NY*: R. Graham Barr, MD, DrPH; John Austin, MD; Belinda D’Souza, MD; Byron Thomashow, MD

*Duke University Medical Center, Durham, NC*: Neil MacIntyre, Jr., MD; H. Page McAdams, MD; Lacey Washington, MD

*HealthPartners Research Institute, Minneapolis, MN*: Charlene McEvoy, MD, MPH; Joseph Tashjian, MD

*Johns Hopkins University, Baltimore, MD*: Robert Wise, MD; Robert Brown, MD; Nadia N. Hansel, MD, MPH; Karen Horton, MD; Allison Lambert, MD, MHS; Nirupama Putcha, MD, MHS

*Lundquist Institute for Biomedical Innovation at Harbor UCLA Medical Center, Torrance, CA*: Richard Casaburi, PhD, MD; Alessandra Adami, PhD; Matthew Budoff, MD; Hans Fischer, MD; Janos Porszasz, MD, PhD; Harry Rossiter, PhD; William Stringer, MD

*Michael E. DeBakey VAMC, Houston, TX:* Amir Sharafkhaneh, MD, PhD; Charlie Lan, DO

*Minneapolis VA*: Christine Wendt, MD; Brian Bell, MD; Ken M. Kunisaki, MD, MS

*Morehouse School of Medicine, Atlanta, GA*: Eric L. Flenaugh, MD; Hirut Gebrekristos, PhD; Mario Ponce, MD; Silanath Terpenning, MD; Gloria Westney, MD, MS

*National Jewish Health, Denver, CO*: Russell Bowler, MD, PhD; David A. Lynch, MB

*Reliant Medical Group, Worcester, MA*: Richard Rosiello, MD; David Pace, MD

*Temple University, Philadelphia, PA*: Gerard Criner, MD; David Ciccolella, MD; Francis Cordova, MD; Chandra Dass, MD; Gilbert D’Alonzo, DO; Parag Desai, MD; Michael Jacobs, PharmD; Steven Kelsen, MD, PhD; Victor Kim, MD; A. James Mamary, MD; Nathaniel Version Date: March 26, 2021 Marchetti, DO; Aditi Satti, MD; Kartik Shenoy, MD; Robert M. Steiner, MD; Alex Swift, MD; Irene Swift, MD; Maria Elena Vega-Sanchez, MD

*University of Alabama, Birmingham, AL*: Mark Dransfield, MD; William Bailey, MD; Surya P. Bhatt, MD; Anand Iyer, MD; Hrudaya Nath, MD; J. Michael Wells, MD

*University of California, San Diego, CA*: Douglas Conrad, MD; Xavier Soler, MD, PhD; Andrew Yen, MD

*University of Iowa, Iowa City, IA*: Alejandro P. Comellas, MD; Karin F. Hoth, PhD; John Newell, Jr., MD; Brad Thompson, MD

*University of Michigan, Ann Arbor, MI*: MeiLan K. Han, MD MS; Ella Kazerooni, MD MS; Wassim Labaki, MD MS; Craig Galban, PhD; Dharshan Vummidi, MD

*University of Minnesota, Minneapolis, MN*: Joanne Billings, MD; Abbie Begnaud, MD; Tadashi Allen, MD

*University of Pittsburgh, Pittsburgh, PA*: Frank Sciurba, MD; Jessica Bon, MD; Divay Chandra, MD, MSc; Joel Weissfeld, MD, MPH

*University of Texas Health, San Antonio, San Antonio, TX*: Antonio Anzueto, MD; Sandra Adams, MD; Diego Maselli-Caceres, MD; Mario E. Ruiz, MD; Harjinder Singh

## Data and Code Availability

LTRC genotyping data are available on dbGaP with accession number phs001662.v1.p1

LTRC RNA-seq data are available through TOPMed, https://topmed.nhlbi.nih.gov. COPDGene data are available on dbGaP with accession numbers phs000179 and phs000765.

