## Supplementary for "Overlap between COPD genetic association results and transcriptional quantitative trait loci"

**Supplementary Methods: Long Read Sequencing**

*cDNA synthesis*

100 ng of RNA was mixed with dNTP and RNA Inhibitor, then denatured to remove secondary structure for 3 minutes at 72C. Prior to reverse transcription (RT), indexed oligo(dT) primers were added to the mix at 50C. First strand synthesis using Maxima H(-) reverse transcriptase (Thermo Fisher) and a template switching oligo was performed for 90 minutes at 50C. The cDNA was then amplified using PCR with SeqAmp polymerase, ISPCR primer, and Nextera A index primer (95C for 1 minute, 15 cycles of 98C for 15 seconds, 65C for 30 seconds, 68C for 6 minutes, 72C for 7 minutes, 4C hold).  Each sample was indexed using a unique combination of olgo(dT) primer and Nextera A index primer. cDNA was cleaned up using ProNex Size Selective Purification System (Promega) at a 1:1.2 ratio.

*Enrichment*

After quantification by Qubit, equal amounts cDNA from every sample were pooled together. 3 ug of cDNA pool were target enriched using the xGEN Hybridization Capture (Integrated DNA Technologies). xGen Human Cot DNA and Blocking Oligos were added to the cDNA pool and cleaned using SPRI beads at a 1.8:1 ratio. The beads were eluted in a mix of xGen 2X Hybridization Buffer, xGen Hybridization Buffer Enhancer, and xGen Hyb Panel and incubated at room temperature for 5 min. The beads were removed using a magnet, and the solution incubated at 95C for 30 seconds, 65C for 4 hours, and a 65C hold. Streptavidin beads were added and incubated at 65C for 45 minutes. The mixture was washed using Wash Buffer 1 and Stringent Wash Buffer at 65C. Then, Wash Buffer 1, Wash Buffer 2, and Wash Buffer 3 were used to wash the mixture at room temperature. PCR was performed on the final solution with SeqAmp polymerase, ISPCR primer, and Nextera A Universal primer for 15 cycles (95C for 1 minute, 98C for 15 seconds, 65C for 30 seconds, 68C for 6 minutes, 72C for 7 minutes, 4C hold). The final enriched library was cleaned up using SPRI beads at 0.65:1 ratio.

*R2C2 library preparation and sequencing*

DNA splints were generated through primer extension as previously described using 2 complementary DNA oligos. Enriched cDNA libraries were circularized by Gibson assembly (NEBbuilder HiFi) with a  DNA splint with ends complementary to the cDNA. ExoI, ExoIII, and Lambda Exonuclease (NEB) were added and incubated for 16 hours at 37C to remove non-circular molecules followed by heat inactivation for 20 minutes at 80C. The reaction was then cleaned using SPRI beads at a 0.8:1 ratio. The clean, circularized library was used as the template for rolling circle amplification (RCA) using Phi29 (NEB) with a random hexamer primer for 18 hours at 30C, and then heat inactivated for 10 minutes at 65C. The reaction was then debranched using T7 endonuclease (NEB) for 2 hours at 37C before being cleaned and concentrated using a Monarch PCR and DNA cleanup kit (NEB).The resulting R2C2 DNA was then run on a 1% agarose gel and DNA  >10kb was excised and purified using a Monarch DNA Gel extraction kit (NEB).

Following the manufacturer’s protocol, we then used the Genomic DNA by Ligation (SQK-LS114) kit from ONT to generate sequencing libraries from this size-selected R2C2 DNA. The final library was loaded onto a PromethION R10.4 flow cell and run at 400bp/s. Approximately once per day, flow cells were flushed and treated with DNAse I, then loaded with additional library to increase sequencing throughput.

Resulting raw reads were basecalled using the SUP model of guppy (v6) and consensus called and demultiplexed using C3POa (v2.3).

Oligos used:

```
cDNA synthesis
ISPCR primer 	AAGCAGTGGTATCAACGCAGAGTAC
Oligo_dT_Index1	AAGCAGTGGTATCAACGCAGAGT [10nt index] ACTTTTTTTTTTTTTTTTTTTTTTTTTTTTTTVN
TSO1_Nextera	TCGTCGGCAGCGTCAGATGTGTATAAGAGArCAG  rUGA  ArU  rUC  TGGTrGrGrG
Nextera_Primer_A1	AATGATACGGCGACCACCGAGATCTACAC  [8nt index]  TCGTCGGCAGCGTCAGATG

Enrichment

NextA_F_Blocking	AATGATACGGCGACCACCGAGATCTACAC  IIIIIIII  TCGTCGGCAGCGTCAGATGTGTATAAGAGACAG/3ddC/
NextA_RC_Blocking	CTGTCTCTTATACACATCTGACGCTGCCGACGA  IIIIIIII  GTGTAGATCTCGGTGGTCGCCGTATCATT
RC_OligodT		AA AAA AAA AAA AAA AAA AAA AAA AAA AAA AGT III III III IAC TCT GCG TTG ATA CCA CTG CTT
IndexedOligodTBlocker	AAG CAG TGG TAT CAA CGC AGA GTI III III III ACT TTT TTT TTT TTT TTT TTT TTT TTT TTT TT/3ddC/

Splint Synthesis

Nextera_A_Splint_1_F	GATCTCGGTGGTCGCCGTATCATT TGAGGCTGATGAGTTCCATA NNNNNTATATNNNNN ATCACTACTTAGTTTTTTGATAGCTTCAAGCCAGAGTTGTCTTTTTCTCTTTGCTGGCAGTAAAAG
ISPCR_Splint_1_R	ACTCTGCGTTGATACCACTGCTT AAAGGGATATTTTCGATCGC NNNNNATATANNNNN TTAGTGCATTTGATCCTTTTACTCCTCCTAAAGAACAACCTGACCCAGCAAAAGGTACACAATACTTTTACTGCCAGCAAAGAG
Nextera_A_Splint_2_F	GATCTCGGTGGTCGCCGTATCATT TGCCGGTTGGGTATCAATAA NNNNNTATATNNNNN ATTGCCTTTATTCTATCTACTTAGTTTTGGCGATGTAGTCTACCTATCCTGATGCTGAATAAAGGC
ISPCR_Splint_2_R	ACTCTGCGTTGATACCACTGCTT AATTAGGTTCTAGGATCACG NNNNNATATANNNNN CTGCCATCGAAAATTTTTCACCCGTAACAAGAACTTACAACTCTCTGACGCCTATATCATGAAGGCCTTTATTCAGCATCAGGA
Nextera_A_Splint_3_F	GATCTCGGTGGTCGCCGTATCATT GTCGTGATCAAACATTGGGT NNNNNTATATNNNNN TAAAAGTTTTCTGTGTCCATTACGTTTTTTGGAGACGGTCTCAACTATTCTTAATCTCGGCGAACT
ISPCR_Splint_3_R	ACTCTGCGTTGATACCACTGCTT AAGTGAGTCGTATTACAGCT NNNNNATATANNNNN ATATTCAGTTCGAAACCTTACCTCCTCTGAAAAGTGTCCCCGAAATCTCTAGGAAATTAACCCAAGTTCGCCGAGATTAAGAAT
Nextera_A_Splint_4_F	GATCTCGGTGGTCGCCGTATCATT TAGTGTAAGGTAGCATCCGT NNNNNTATATNNNNN TACGATCATATTATGTTCGTCTGTCTTTAACGGGCTATACTTCTTCTTTAGGAAAGGTCTGAACCT
ISPCR_Splint_4_R	ACTCTGCGTTGATACCACTGCTT CTACTATTTGAGTAAAGGCG NNNNNATATANNNNN ACCTGTGTATTAGACCCCACTACTACCCTTAAGATTCCATAACCTTAAGTAGAACCCAGATTGAAGGTTCAGACCTTTCCTAAA
```

**Supplementary Results:**

*Identification of an additional colocalization between FBXO38 splicing and COPD GWAS*

We found that rs10037493-T (5:148475407:C:T), which is associated with increased COPD risk is also associated with decreased inclusion of the identified exon (Supplementary Figure 1 a-b). This variant has a 71% probability of being causal for the sQTL association according to SuSie finemapping [23]. While long read sequencing did not identify a confirmed isoform containing this exon (ie in the isoforms passing filtering thresholds), in the raw RNAseq reads we identified at least 15 reads supporting the presence of this exon (Supplementary Figure 1c and Supplementary Figure 2). These reads correspond to two predicted protein-coding isoforms, XM_017009902 and XM_011537684, which would code for 713 and 788 amino acid proteins which differ from the usual 1188 amino acid protein.

**Supplementary Table 1**

| Locus (GRCh38) | Locus | Top associated GWAS  variant | Best colocalized Gene | Best Colocalized Type | Other colocalized genes | GWAS identified target gene |
| --- | --- | --- | --- | --- | --- | --- |
| 4:144567946:A:G | 4q31.21 | rs13140176 | HHIP | LTRC_sQTL | LTRC_eQTL_HHIP_0.9 | HHIP |
| 4:105897896:G:A | 4q24 | rs34712979 | NPNT | LTRC_sQTL |  | PPA2, NPNT |
| 5:148475407:C:T | 5q32 | rs10037493 | FBXO38 | LTRC_sQTL | COPDGene_sQTL_ABLIM3_0.89 | FBXO38 |
| 15:71329185:G:A | 15q23 | rs1441358 | THSD4 | LTRC_eQTL | LTRC_eQTL_THSD4_0.99 | THSD4, LARP6 |
| 16:75439564:G:A | 16q23.1 | rs4888379 | TMEM170A | COPDGene_sQTL | COPDGene_sQTL_CFDP1_0.8 | CFDP1, TERF2IP, CTRB2 |
| 6:30814428:G:C | 6p21.33 | rs2284174 | IER3 | COPDGene_sQTL | LTRC_sQTL_FLOT1_0.97; LTRC_sQTL_PPT2-EGFL8_0.85; LTRC_sQTL_NRM_0.8 | IER3, FLOT1, TUBB, DDR1, VARS2 ,SFTA2, CDSN |
| 4:88948181:T:C | 4q22.1 | rs7671261 | PKD2 | COPDGene_sQTL |  | FAM13A, NAP1L5,PPM1K |
| 6:32660630:T:C | 6p21.32 | rs2070600 | . | COPDGene_sQTL | LTRC_eQTL_AGER_0.99; LTRC_sQTL_PSORS1C1_0.95; COPDGene_sQTL_CCHCR1_0.95; COPDGene_eQTL_VARS2_0.94; COPDGene_sQTL_VARS2_0.94; LTRC_sQTL_NOTCH4_0.93; LTRC_sQTL_LST1_0.93; LTRC_sQTL_AIF1_0.92; COPDGene_sQTL_AGER_0.9; LTRC_sQTL_CCHCR1_0.88; LTRC_sQTL_SFTA2_0.86; LTRC_sQTL_VARS2_0.8 | AGER, NOTCH4, AGPAT1, CYP21A2, MSH5, LY6G6E  HLA-DQB1 |
| 5:157505976:A:T | 5q33.3 | rs10866659 | ADAM19 | LTRC_eQTL | COPDGene_sQTL_ADAM19_0.89 | ADAM19, CYFIP2, CLINT1, SOX30, C5orf52 |
| 3:128242335:T:A | 3q21.3 | rs2955083 | EEFSEC | COPDGene_sQTL | COPDGene_sQTL_RUVBL1_0.91 | RUVBL1, GATA2 |
| 2:238965524:G:A | 2q37.3 | rs62191105 | TWIST2 | LTRC_eQTL |  | UBE2F |
| 3:25496178:G:A | 3p24.2 | rs1529672 | RARB | LTRC_eQTL |  | RARB |
| 2:9145396:G:A | 2p25.1 | rs955277 | LINC00299 | COPDGene_sQTL |  | ASAP2 |
| 1:16979534:C:A | 1p36.13 | rs9435731 | CROCC | COPDGene_sQTL | LTRC_sQTL_MST1P2_0.98; LTRC_eQTL_ATP13A2_0.98; LTRC_sQTL_CROCC_0.98; LTRC_sQTL_MFAP2_0.98; LTRC_sQTL_CROCCP2_0.97; LTRC_eQTL_MFAP2_0.96; LTRC_APAQTL_MFAP2_0.82 | MFAP2  PADI2 |
| 7:100032719:C:T | 7q22.1 | rs2897075 | ZSCAN21 | COPDGene_sQTL | COPDGene_eQTL_MCM7_0.98; COPDGene_APAQTL_ZKSCAN1_0.97; COPDGene_APAQTL_STAG3L5P_0.96; COPDGene_sQTL_CNPY4_0.95; LTRC_sQTL_COPS6_0.93; LTRC_sQTL_ZSCAN21_0.8 | ZKSCAN1, ZNF3, AZGP1, AP4M1, CNPY4, TRIM4, PVRIG, NYAP1 |
| 19:45790878:G:A | 19q13.32 | rs72626215 | DMWD | LTRC_APAQTL | LTRC_APAQTL_SYMPK_0.97; LTRC_sQTL_QPCTL_0.82 | DMPK, SNRPD2 |
| 1:39582337:G:A | 1p34.3 | rs76841360 | PPIEL | COPDGene_sQTL | COPDGene_sQTL_MACF1_0.88; COPDGene_sQTL_PABPC4_0.85 | PABPC4, OXCT2, MYCL |
| 12:95843792:T:C | 12q23.1 | rs7307510 | NTN4 | LTRC_sQTL | COPDGene_sQTL_CFAP54_0.88 | AMDHD1 |
| 4:74748514:C:T | 4q13.3 | rs4585380 | BTC | LTRC_sQTL |  | EPGN |
| 1:111195294:T:C | 1p13.3 | rs629619 | DENND2D | COPDGene_sQTL | LTRC_eQTL_KCNA2_0.84 | DRAM2, OVGP1, INKA2 |
| 14:92649065:G:C | 14q32.12 | rs72699855 | RIN3 | LTRC_APAQTL |  | CPSF2, RIN3, ITPK1 |
| 1:239689643:G:C | 1q43 | rs11579382 | CHRM3-AS2 | COPDGene  APAQTL | COPDGene_sQTL_CHRM3-AS2_0.95 |  |
| 11:13145018:T:C | 11p15.2 | rs4757118 | RASSF10 | LTRC_eQTL |  | PARVA |
| 5:151215512:A:G | 5q33.1 | rs979453 | GM2A | LTRC_APAQTL | COPDGene_APAQTL_GM2A_0.95 |  |
| 7:2830820:T:G | 7p22.3 | rs798565 | GNA12 | LTRC_sQTL |  | GNA12 |
| 17:38730575:G:A | 17q12 | rs34727469 | CISD3 | LTRC_eQTL | LTRC_eQTL_PCGF2_0.94; COPDGene_eQTL_MLLT6_0.94; LTRC_eQTL_EPOP_0.92; COPDGene_sQTL_MLLT6_0.9; COPDGene_eQTL_AC006449.6_0.88 | CACNB1 |
| 3:29430921:G:C | 3p24.1 | rs13073544 | RBMS3 | LTRC_eQTL |  | RBMS3 |
| 15:49578340:A:G | 15q21.2 | rs72731149 | FAM227B | LTRC_sQTL | COPDGene_sQTL_FAM227B_0.92; COPDGene_sQTL_AP4E1_0.9 | FAM227B, SLC27A2 |
| 16:58063696:G:C | 16q21 | rs8044657 | MMP15 | LTRC_sQTL |  | CNGB1 |
| 10:80458350:G:C | 10q22.3 | rs721917 | TSPAN14 | COPDGene_sQTL | COPDGene_sQTL_FAM213A_0.95; LTRC_APAQTL_SFTPA2_0.89; LTRC_sQTL_DYDC2_0.89 | TMEM254, DYDC2, FAM213A |
| 1:45543641:G:C | 1p34.1 | rs4660861 | TESK2 | COPDGene  APAQTL | COPDGene_eQTL_AKR1A1_0.95; COPDGene_sQTL_AKR1A1_0.95; COPDGene_APAQTL_MUTYH_0.93; LTRC_sQTL_AKR1A1_0.87 | MUTYH, IPP, UROD |
| 2:42206107:C:T | 2p21 | rs12466981 | COX7A2L | COPDGene  APAQTL |  | EML4 |
| 17:30129740:C:T | 17q11.2 | rs8080772 | NSRP1 | COPDGene_sQTL | LTRC_sQTL_NSRP1_0.82 | SLC6A4, EFCAB5, GOSR1  CORO6 |

**Supplementary Figures**

Supplementary Figure 1

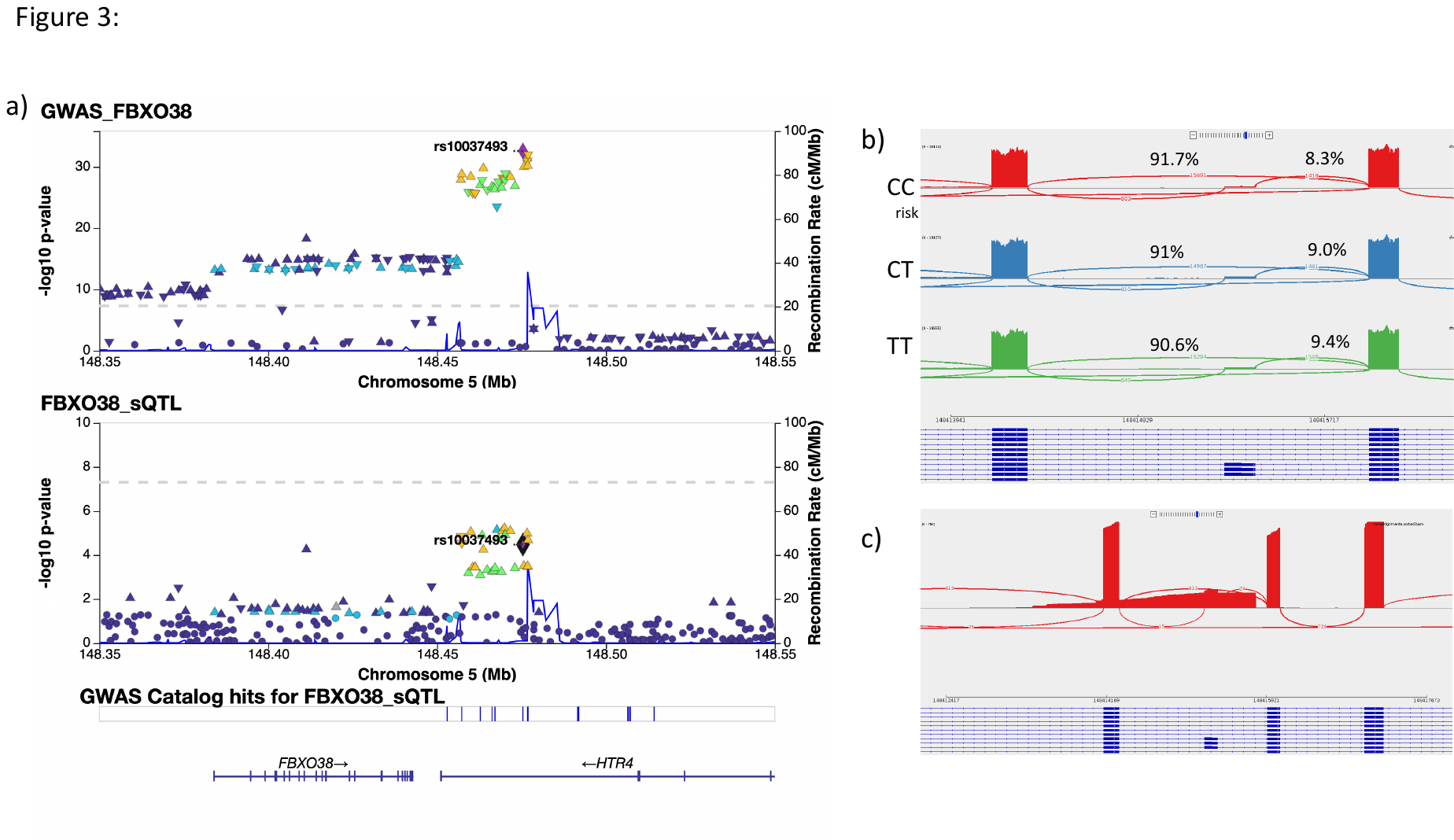

**Supplementary Figure 1: An additional colocalization between *FBXO38* sQTLs and COPD.** a) Locus association plot for COPD GWAS and *FBXO38* sQTL. The lead colocalized SNP, rs10037493, is highlighted in purple and used as the LD reference. b) IGV sashimi plot showing the region spanning chr5:148413941-148416605 for 273 subjects from each genotype of rs10037493. c) Long read sequencing data showing raw FBXO38 reads of isoforms containing the exon of interest.

Supplementary Figure 2

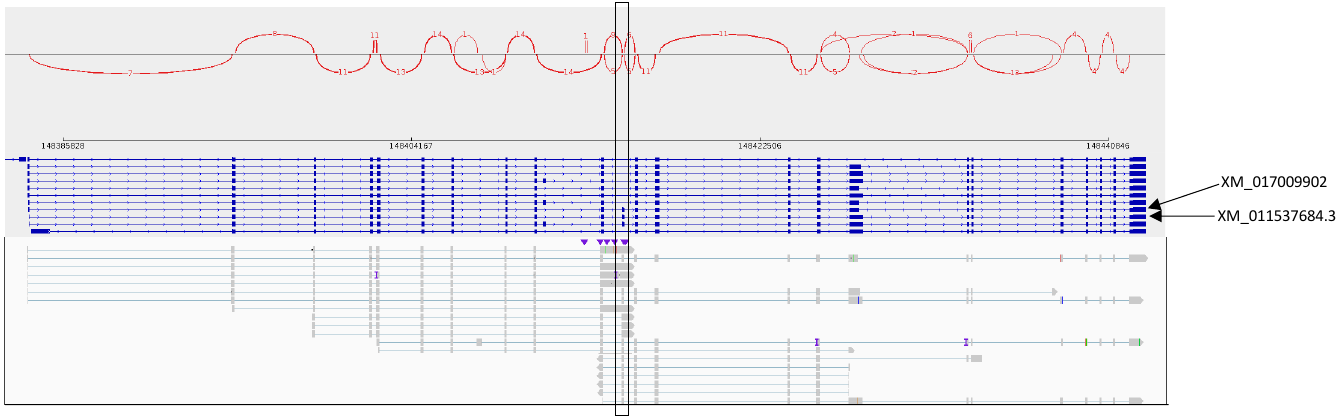

**Supplementary Figure 2: IGV plot of raw long read sequencing data from FBXO38 showing reads containing the exon**
